# Initiation of SGLT2 inhibitors among adults with type 2 diabetes and cardiorenal disease in the All of Us Research Program, 2019–2023

**DOI:** 10.64898/2026.11.19.26368468

**Authors:** Merrera Kebeba, Luboom Taa, Getahun Alemu, Kaleb Lema, Lidetu Kayamo, Chala Oljira, Kemal Awol

## Abstract

**Background:** Sodium-glucose cotransporter 2 (SGLT2) inhibitors reduce cardiovascular and kidney events in adults with type 2 diabetes(T2DM) and cardiorenal disease, but treatment initiation among eligible patients remains incompletely characterized. We estimated the cumulative incidence of recorded SGLT2 inhibitor initiation and associated participant characteristics in a diverse U.S. cohort.

**Methods:** We conducted a retrospective cohort study leveraging electronic health record data from the All of Us Research Program. Adults with confirmed type 2 diabetes, at least one co-occurring cardiorenal condition (atherosclerotic cardiovascular disease, heart failure, or chronic kidney disease), and no prior SGLT2 inhibitor use were followed from the earliest date all criteria were met during 2019–2023. The 365-day cumulative incidence of initiation was estimated using the Aalen–Johansen method, accounting for competing risk of all-cause mortality. Cause-specific modified Poisson regression with robust error variance yielded adjusted risk ratios (aRRs) for initiation predictors

**Results:** Among 9,333 eligible participants, 637 had recorded SGLT2 inhibitor initiation within 365 days (estimated cumulative incidence, 6.85% (95% CI 6.34–7.36)). The 365-day incidence was 8.40% among individuals with heart failure, 6.65% with atherosclerotic cardiovascular disease, and 5.28% with chronic kidney disease. Initiation was more likely with heart failure (RR 1.51, 95% CI 1.29–1.76), and glycated hemoglobin ≥9.0% versus <7.0% (RR 2.61, 95% CI 2.07–3.28), but less likely with chronic kidney disease (RR 0.63, 95% CI 0.54–0.74), age ≥75 versus 45–64 years (RR 0.51, 95% CI 0.40–0.65), and non-Hispanic Black versus non-Hispanic White race and ethnicity (RR 0.79, 95% CI 0.65–0.97).

**Conclusions:** Fewer than 1 in 10 eligible, untreated adults with T2D and cardiorenal disease initiated SGLT2 inhibitor therapy within one year. Even with expanding guideline indications, substantial implementation gaps persist, particularly among patients with CKD, older adults, and non-Hispanic Black adults. Targeted health systems are imperative to optimize the use of guideline-recommended cardiorenal therapies.

## Introduction

Sodium-glucose cotransporter 2 (SGLT2) inhibitors reduce hospitalization for heart failure, kidney disease progression, and major adverse cardiovascular events in adults with type 2 diabetes mellitus (T2DM) [1]. Since 2019, systematic reviews and meta-analyses of clinical trials, alongside real-world evidence, have demonstrated benefits in heart failure across ejection fractions [2] and in chronic kidney disease (CKD), including in patients without diabetes [3]. Over the same period, clinical guidelines across endocrinology, cardiology, and nephrology increasingly recommended SGLT2 inhibitors for adults with T2DM and atherosclerotic cardiovascular disease (ASCVD), heart failure, or CKD [4–8]. The 2022 American Diabetes Association (ADA) and European Association for the Study of Diabetes (EASD) consensus report recommended these agents for cardiorenal benefit regardless of baseline glycated hemoglobin (HbA1c) or metformin use [7], consistent with current guidelines [9–12].

Translation of this evidence into practice has been slow and uneven. In the first years of SGLT2 inhibitor approval, initiation was less likely among older adults and patients with CKD [13], and use was lower among Black and female patients and those with lower household income [14]. Across 28 U.S. health systems in 2022–2023, 11.9% of adults with diabetes who met Class 1A recommendation criteria received an SGLT2 inhibitor prescription, and no health system exceeded 25% prescribing rate [15]. In the Veterans Health Administration, 14.6% of patients with T2DM, heart failure, and ASCVD had recorded use in 2020 [16], and in a national registry, 20.2% of eligible patients hospitalized for heart failure with reduced ejection fraction received a discharge prescription for an SGLT2 inhibitor in 2021–2022[17]. Utilization was also low among patients with CKD, particularly those with more advanced disease [18,19]. In the All of Us Research Program, recorded SGLT2 inhibitor use was 8.6% among participants with diabetes and ASCVD and below 10% among those with diabetes and heart failure or CKD [20], and the prescription of newer glucose-lowering agents differed by race and ethnicity [21].

Most published estimates describe the prevalence of SGLT2 inhibitor use rather than initiation during follow-up among eligible individuals without prior exposure. One U.S. electronic health record (EHR) study examined new prescriptions after an incident ASCVD diagnosis [22], but longitudinal evidence spanning ASCVD, heart failure, and CKD in diverse populations remains limited. Using EHR data from the All of Us Research Program, we estimated the 1-year cumulative incidence of first recorded SGLT2 inhibitor initiation among adults with T2DM and ASCVD, heart failure, or CKD without prior recorded use, and examined variation by the calendar year of cohort entry (2019–2023), cardiorenal condition, and participant characteristics.

## Methods

### Study design and data source

We conducted a retrospective cohort study using de-identified data from the All of Us Research Program (AoU), a nationwide initiative designed to advance precision medicine through the integration of multidimensional health data. The program recruits adults aged 18 years and older from more than 400 enrollment sites across the United States, particularly populations historically underrepresented in biomedical research [23,24]. AoU links participant-provided longitudinal data (including surveys, standardized physical measurements, biospecimens, and wearables) with electronic health record (EHR) data from participating healthcare organizations, which are harmonized to the Observational Medical Outcomes Partnership (OMOP) common data model [25,26]. All participants provided written informed consent for research participation, including consent for long-term follow-up through linkage to their EHRs.

We used data from the AoU Controlled Tier Curated Data Repository version 9 (CDR v9; dataset release C2025Q4R6, released June 30, 2026), which contains data from 747,029 adult participants enrolled between May 31, 2017, and January 1, 2025 [27]. We assessed cohort entry from January 1, 2019, through December 31, 2023, and ascertained outcomes through December 31, 2024. Reporting follows the Strengthening the Reporting of Observational Studies in Epidemiology (STROBE) statement and its Reporting of Studies Conducted Using Observational Routinely Collected Health Data (RECORD) extension [28,29] (S1 Checklist).

### Cardiorenal phenotypes

We identified T2DM, heart failure, CKD, and ASCVD from structured EHR diagnosis and procedure records using clinically reviewed OMOP concept sets. For T2DM, heart failure, and CKD, we adapted the Centers for Medicare & Medicaid Services Chronic Conditions Warehouse algorithms [30], requiring either one qualifying inpatient diagnosis or two outpatient-type diagnoses (outpatient, office, telehealth, or emergency department encounters) on separate dates within 730 days. ASCVD was defined by an inpatient diagnosis of myocardial infarction or ischemic stroke, a coronary revascularization procedure (coronary artery bypass grafting or percutaneous coronary intervention), or two outpatient-type diagnoses of coronary or atherosclerotic peripheral arterial disease on separate dates within 730 days. Each condition was dated to the first date its criteria were met. Operational definitions are provided in the supplementary material (Tables A S1 Appendix).

### Eligibility and index date

Participants were eligible if they were aged ≥18 years, had confirmed T2DM and at least one confirmed cardiorenal condition (ASCVD, heart failure, or CKD), and had at least 365 days of prior EHR history with at least one clinical encounter in each half of the preceding year. Eligibility was assessed throughout 2019–2023, and the index date was defined as the earliest date on which all inclusion criteria were met. Exclusion criteria included a recorded death on or before cohort entry, documented pregnancy at baseline, maintenance dialysis in the preceding 365 days, prior kidney transplantation, and prior recorded SGLT2 inhibitor use. Furthermore, participants were excluded if their most recent estimated glomerular filtration rate (eGFR) in the preceding 365 days was below 20 mL/min/1.73 m², consistent with the 2022 Kidney Disease: Improving Global Outcomes (KDIGO) guidelines [8]; however, individuals without a recorded eGFR were retained (Fig. 1).

**Fig 1.**
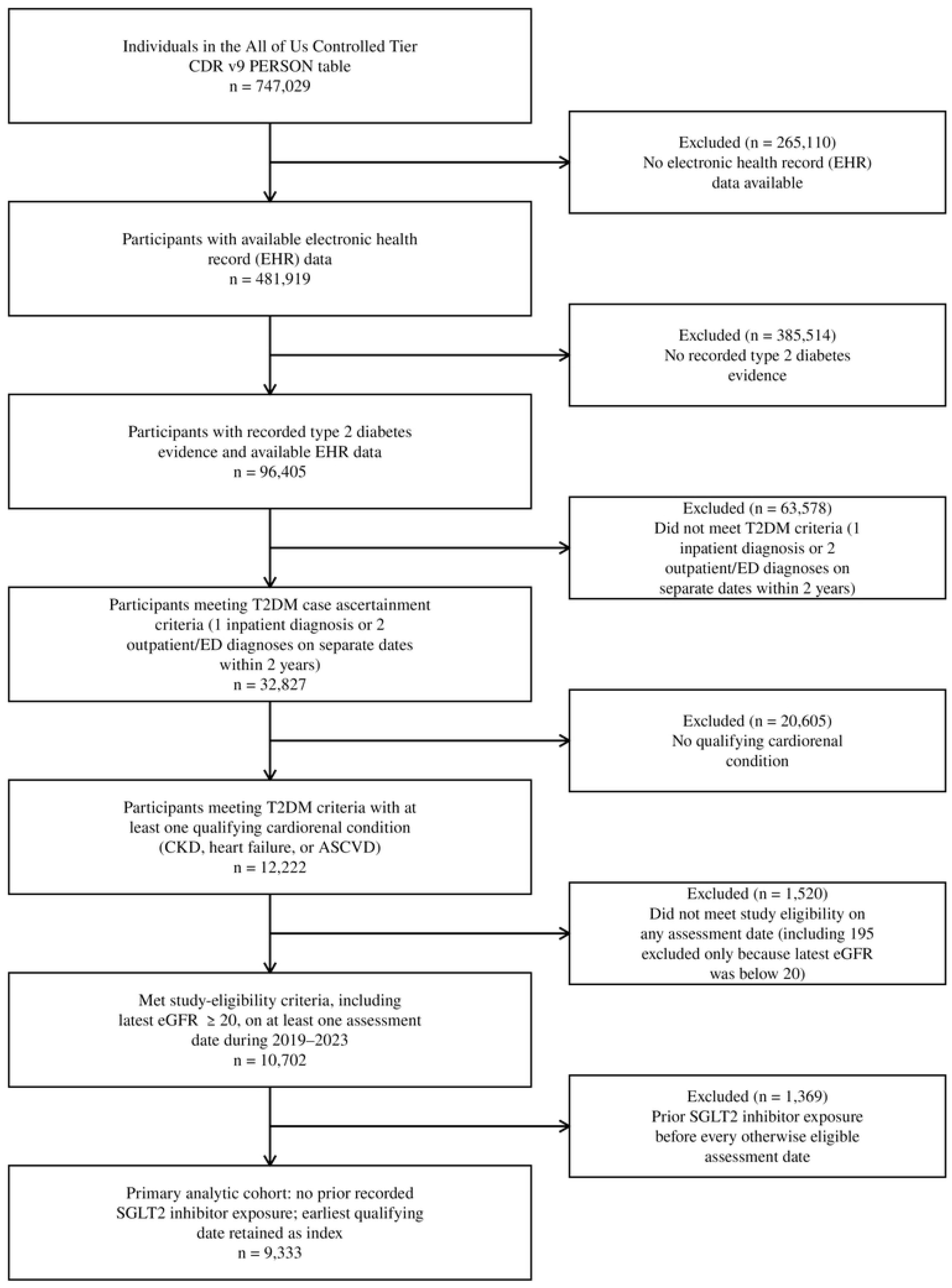
Flow diagram of cohort selection. Counts are unique participants. Eligibility was assessed on every date from 2019 through 2023, and each participant’s earliest eligible date without prior recorded SGLT2 inhibitor exposure was retained as the index date. ASCVD, atherosclerotic cardiovascular disease; CKD, chronic kidney disease; eGFR, estimated glomerular filtration rate; SGLT2, sodium-glucose cotransporter 2; T2DM, type 2 diabetes mellitus.

### SGLT2 inhibitor ascertainment and outcomes

SGLT2 inhibitor records were identified in the drug exposure domain using RxNorm and RxNorm Extension concepts for bexagliflozin, canagliflozin, dapagliflozin, empagliflozin, ertugliflozin, and sotagliflozin, including single-ingredient and combination products. Records originating from research activities or participant surveys were excluded.

The primary outcome was the first recorded SGLT2 inhibitor initiation within 365 days of the index date; initiation within 180 days was a secondary outcome. Follow-up ended at initiation, death, the end of EHR observation, the outcome horizon, or December 31, 2024, whichever occurred first.

### Covariates

Demographic characteristics included age, participant-reported sex at birth, and race and ethnicity. Baseline clinical measures, including estimated glomerular filtration rate (eGFR), HbA1c, urine albumin-to-creatinine ratio (UACR), body mass index (BMI), and blood pressure, were based on the most recent valid measurement in the 365 days before the index date, unless otherwise specified (S1 Appendix). eGFR was calculated from serum creatinine using the 2021 race-free Chronic Kidney Disease Epidemiology Collaboration equation [31]; if creatinine was unavailable on the measurement date, the laboratory-reported eGFR was used. Among participants with heart failure, left ventricular ejection fraction (LVEF) was categorized from the most recent pre-index measurement. Baseline exposure to glucose-lowering and cardiovascular medications was ascertained from EHR medication records in the preceding year. Health care utilization was assessed by the number of distinct outpatient contact days and any inpatient or emergency department use. Insurance status and difficulty affording prescription medicines were derived from participant surveys completed at any point during program enrollment (Table F in S1 Appendix).

### Statistical analysis

Baseline characteristics were summarized overall and by 365-day initiation status as counts with percentages or means with standard deviations (SDs), and differences were described with absolute standardized mean differences [32]. Participants whose follow-up ended before day 365 without an event were included only in the overall baseline summary. Cumulative incidence of recorded initiation and of death before initiation was estimated with the Aalen–Johansen estimator, treating death as a competing event [33,34]. Estimates were obtained overall at 365 and 180 days and by index year and cardiorenal condition. Because the ASCVD, heart failure, and CKD groups overlap, condition-specific estimates were descriptive and were not compared formally. Percentile 95% confidence intervals (CIs) were obtained from 2,000 participant-level bootstrap samples.

Associations between participant characteristics and initiation within 365 days were estimated as risk ratios (RRs) with modified Poisson regression [35,36]. Each characteristic was examined in a separate unadjusted model and in a multivariable model adjusted for index year, age (natural cubic spline), sex at birth, and race and ethnicity. Death before initiation was classified as no initiation, and participants censored without an event before day 365 were excluded from these models. Missing clinical values were analyzed as explicit not-measured categories without imputation, and participants with unknown sex at birth or race and ethnicity were excluded.

### Sensitivity analyses

The cohort was rebuilt from eligibility onward with calendar-specific eGFR thresholds of 30 mL/min/1.73 m² in 2019–2020, 25 in 2021–2022, and 20 in 2023, reflecting evolving guidance [5,8]. Associations were re-estimated in a model including all characteristics simultaneously, with age modeled continuously, under the following variations:

- Application of inverse probability of censoring weights for participants censored before day 365.
- Application of a cause-specific proportional hazards model.
- Exclusion of index-day medication records(may reflect medication reconciliation).
- Use of 180-day outcome window.
- Restriction to participants with any baseline medication record.
- Exclusion of participants entering in the fourth quarter of 2023.

### Data protection

In accordance with the All of Us Data and Statistics Dissemination Policy, counts of 1–20 participants and statistics from which such counts could be derived are not reported and are shown as NR.

### Ethics statement

The All of Us Research Program protocol and consent materials were approved by the All of Us Institutional Review Board (IRB), and participants consented to participation and secondary research use of their data. The All of Us IRB determined that secondary research using Controlled Tier data in the Researcher Workbench does not constitute human subjects research under 45 CFR 46 and does not require project-specific review (protocol 2021-02-TN-001). Analyses were conducted by authorized researchers in the Researcher Workbench under an institutional data use agreement and the All of Us Data User Code of Conduct. Data were accessed between July 20, 2026, and November 1, 2026.

### Data Availability Statement

The data underlying this study are available to authorized researchers through the NIH All of Us Researcher Workbench. Participant-level data cannot be publicly shared due to privacy restrictions. Access to the Controlled Tier Curated Data Repository (CDR v9, C2025Q4R6) requires appropriate authorization under All of Us policies.

## Results

### Study cohort

Of the 481,919 individuals with electronic health record (EHR) data in the All of Us Controlled Tier CDR v9, 96,405 had an EHR diagnosis of T2DM, 32,827 met the T2DM confirmation criteria, and 12,222 also had confirmed ASCVD, heart failure, or CKD (Fig 1; Tables C and D in S1 Appendix). Of these, 9,333 met all eligibility criteria on at least one date during 2019–2023, with 53.5% entering the cohort in 2019 (Table 1). The mean age was 64.2 years (SD 11.4) and 50.9% reported female sex at birth. 45.1% were non-Hispanic White, 25.9% were non-Hispanic Black, and 18.2% were Hispanic or Latino. ASCVD, heart failure, and CKD were present in 48.8%, 28.7%, and 50.3% of participants, respectively (Table C in S1 Appendix). The median follow-up was 365 days, yielding 8,872 person-years of observation.

**Table 1.** Baseline characteristics of the analytic cohort, overall and by recorded SGLT2 inhibitor initiation wthin 365 days.

| **Characteristic** | **Overall (n = 9,333)** | **Initiation by day 365 (n = 637)** | **No initiation by day 365 (n = 8,640)** | **Absolute SMD** |
|---|---|---|---|---|
| **Age group, years, n (%)** |  |  |  |  |
| 18–44 | 519 (5.6) | 42 (6.6) | 473 (5.5) | 0.047 |
| 45–64 | 3,932 (42.1) | 313 (49.1) | 3,593 (41.6) | 0.152 |
| 65–74 | 3,239 (34.7) | 209 (32.8) | 3,018 (34.9) | 0.045 |
| ≥75 | 1,643 (17.6) | 73 (11.5) | 1,556 (18.0) | 0.186 |
| **Sex at birth, n (%)** |  |  |  |  |
| Female | 4,752 (50.9) | NR | 4,421 (51.2) | 0.047 |
| Male | 4,467 (47.9) | 324 (50.9) | 4,108 (47.5) | 0.066 |
| Other, unknown, or no response | 114 (1.2) | NR | 111 (1.3) | NR |
| **Race and ethnicity, n (%)** |  |  |  |  |
| Non-Hispanic White | 4,206 (45.1) | 292 (45.8) | 3,889 (45.0) | 0.017 |
| Non-Hispanic Black | 2,413 (25.9) | 132 (20.7) | 2,262 (26.2) | 0.129 |
| Hispanic or Latino | 1,702 (18.2) | 141 (22.1) | 1,556 (18.0) | 0.103 |
| Non-Hispanic Asian | 240 (2.6) | NR | 214 (2.5) | 0.090 |
| Other or multiple races/ethnicities | 581 (6.2) | 35 (5.5) | 539 (6.2) | 0.032 |
| Unknown or not reported | 191 (2.0) | NR | 180 (2.1) | NR |
| **Cardiorenal condition, n (%)** |  |  |  |  |
| ASCVD | 4,558 (48.8) | 302 (47.4) | 4,226 (48.9) | 0.030 |
| Heart failure | 2,676 (28.7) | 224 (35.2) | 2,430 (28.1) | 0.152 |
| CKD | 4,690 (50.3) | 247 (38.8) | 4,420 (51.2) | 0.251 |
| **eGFR, mL/min/1.73 m<sup>2</sup>, n (%)** |  |  |  |  |
| ≥60 | 4,468 (47.9) | 342 (53.7) | 4,099 (47.4) | 0.125 |
| 45–59 | 1,738 (18.6) | 124 (19.5) | 1,604 (18.6) | 0.023 |
| 30–44 | 1,038 (11.1) | NR | 958 (11.1) | 0.031 |
| 20–29 | 439 (4.7) | NR | 421 (4.9) | NR |
| Not measured or bounded<sup>a</sup> | 1,650 (17.7) | 82 (12.9) | 1,558 (18.0) | 0.143 |
| **UACR, mg/g, n (%)** |  |  |  |  |
| 20–29 | 995 (10.7) | 92 (14.4) | 900 (10.4) | 0.122 |
| 30–<300 | 538 (5.8) | 53 (8.3) | 480 (5.6) | 0.109 |
| ≥300 | 253 (2.7) | 22 (3.5) | 230 (2.7) | 0.046 |
| Not measured or bounded<sup>a</sup> | 7,547 (80.9) | 470 (73.8) | 7,030 (81.4) | 0.183 |
| **HbA1c category, %, n (%)** |  |  |  |  |
| <7.0 | 3,899 (41.8) | 206 (32.3) | 3,672 (42.5) | 0.211 |
| 7.0–<9.0 | 2,078 (22.3) | 188 (29.5) | 1,880 (21.8) | 0.178 |
| ≥9.0 | 873 (9.4) | 105 (16.5) | 758 (8.8) | 0.234 |
| Not measured | 2,483 (26.6) | 138 (21.7) | 2,330 (27.0) | 0.124 |
| **Body mass index category, kg/m<sup>2</sup>, n (%)** |  |  |  |  |
| <25.0 | 1,058 (11.3) | 48 (7.5) | 998 (11.6) | 0.137 |
| 25.0–29.9 | 2,314 (24.8) | 141 (22.1) | 2,157 (25.0) | 0.067 |
| ≥30.0 | 5,538 (59.3) | 425 (66.7) | 5,088 (58.9) | 0.163 |
| Not measured | 423 (4.5) | 23 (3.6) | 397 (4.6) | 0.050 |
| Systolic blood pressure, mm Hg, mean (SD) | 132 (19) | 133 (20) | 132 (19) | 0.031 |
| Not measured | 1,339 (14.3) | 92 (14.4) | 1,240 (14.4) | 0.003 |
| Diastolic blood pressure, mm Hg, mean (SD) | 75 (12) | 77 (13) | 75 (11) | 0.133 |
| Not measured | 1,339 (14.3) | 92 (14.4) | 1,240 (14.4) | 0.003 |
| **Baseline medication records, n (%)** |  |  |  |  |
| Metformin | 3,214 (34.4) | 331 (52.0) | 2,868 (33.2) | 0.387 |
| Insulin | 3,674 (39.4) | 289 (45.4) | 3,352 (38.8) | 0.133 |
| Sulfonylurea | 1,094 (11.7) | 120 (18.8) | 969 (11.2) | 0.215 |
| DPP-4 inhibitor | 709 (7.6) | 82 (12.9) | 623 (7.2) | 0.189 |
| GLP-1 RA or tirzepatide | 968 (10.4) | 137 (21.5) | 825 (9.5) | 0.335 |
| Statin | 5,625 (60.3) | 441 (69.2) | 5,149 (59.6) | 0.202 |
| ACE inhibitor, ARB, or ARNI | 5,004 (53.6) | 412 (64.7) | 4,563 (52.8) | 0.243 |
| Mineralocorticoid receptor antagonist | 793 (8.5) | 94 (14.8) | 695 (8.0) | 0.212 |
| **Outpatient contact days, mean (SD)<sup>b</sup>** | 27 (24) | 26 (22) | 27 (24) | 0.004 |
| **Health care use, n (%)** |  |  |  |  |
| Any inpatient or ED use | 4,424 (47.4) | 313 (49.1) | 4,077 (47.2) | 0.039 |
| **Health insurance (any date)<sup>e</sup>, n (%)** |  |  |  |  |
| Reported covered | 8,229 (88.2) | 577 (90.6) | 7,606 (88.0) | 0.083 |
| Explicitly uninsured | NR | NR | NR | NR |
| Other, unknown, or no response | NR | NR | NR | 0.083 |
| **Difficulty affording prescriptions (any date)<sup>e</sup> n (%)** |  |  |  |  |
| No | 3,746 (40.1) | 266 (41.8) | 3,473 (40.2) | 0.032 |
| Yes | 721 (7.7) | 53 (8.3) | 664 (7.7) | 0.023 |
| No response | 4,866 (52.1) | 318 (49.9) | 4,503 (52.1) | 0.044 |
| **Index year, n (%)** |  |  |  |  |
| 2019 | 4,993 (53.5) | 193 (30.3) | 4,781 (55.3) | 0.523 |
| 2020 | 1,106 (11.9) | 70 (11.0) | 1,024 (11.9) | 0.027 |
| 2021 | 1,040 (11.1) | 91 (14.3) | 938 (10.9) | 0.104 |
| 2022 | 1,138 (12.2) | 154 (24.2) | 974 (11.3) | 0.343 |
| 2023 | 1,056 (11.3) | 129 (20.3) | 923 (10.7) | 0.267 |

Values are n (%) unless otherwise indicated. All baseline characteristics are summarized for the full analytic cohort (N = 9,333). Clinical measures, medication records, and health care use were assessed in the 365 days before the index date.
<sup>a</sup> Not measured or reported only as a value bound (for example, “>60”) that did not identify the category.
<sup>b</sup> Outpatient encounter days were defined as distinct calendar days with at least one recorded outpatient visit during the 365 days before the index date.
<sup>e</sup> Survey responses from any date, which may precede or follow the index date.
NR, not reported under the All of Us Data and Statistics Dissemination Policy (counts of 1–20 and values from which they could be derived).
ACE, angiotensin-converting enzyme; ARB, angiotensin receptor blocker; ARNI, angiotensin receptor–neprilysin inhibitor; ASCVD, atherosclerotic cardiovascular disease; CKD, chronic kidney disease; DPP-4, dipeptidyl peptidase-4; ED, emergency department; eGFR, estimated glomerular filtration rate; GLP-1 RA, glucagon-like peptide-1 receptor agonist; HbA1c, glycated hemoglobin; LVEF, left ventricular ejection fraction; SD, standard deviation; SGLT2, sodium-glucose cotransporter 2; SMD, standardized mean difference; UACR, urine albumin-to-creatinine ratio.

### Incidence of SGLT2 inhibitor initiation

Among the 9,333 participants, 637 had a recorded SGLT2 inhibitor initiation within 365 days, 80 died before initiation, 56 were censored without an event, and 8,560 remained alive without recorded initiation. The estimated 365-day cumulative incidence was 6.85% (95% CI 6.34–7.36) for initiation and 0.86% (95% CI 0.68–1.05) for death before initiation (Table 2). At 180 days, the cumulative incidence of initiation was 4.21% (95% CI 3.79–4.63).

**Table 2.** Cumulative incidence of recorded SGLT2 inhibitor initiation at 365 and 180 days, overall, by index year, and by cardiorenal condition.

| **Group** | **Initiations/Participants (n/N)** | **Deaths before initiation, n** | **Cumulative incidence, % (95% CI)** |
|---|---|---|---|
| **365-day horizon** |  |  |  |
| All participants | 637/9,333 | 80 | 6.85 (6.34–7.36) |
| **Index year** |  |  |  |
| 2019 | 193/4,993 | 27 | 3.87 (3.34–4.38) |
| 2020 | 70/1,106 | NR | 6.37 (4.93–7.84) |
| 2021 | 91/1,040 | 22 | 8.79 (7.12–10.57) |
| 2022 | 154/1,138 | NR | 13.59 (11.59–15.61) |
| 2023 | 129/1,056 | NR | 12.25 (10.29–14.23) |
| **Cardiorenal condition<sup>a</sup>** |  |  |  |
| ASCVD | 302/4,558 | 47 | 6.65 (5.90–7.38) |
| Heart failure | 224/2,676 | 46 | 8.40 (7.41–9.53) |
| CKD | 247/4,690 | 53 | 5.28 (4.62–5.92) |
| **180-day horizon** |  |  |  |
| All participants | 392/9,333 | 42 | 4.21 (3.79–4.63) |
| **Index year** |  |  |  |
| 2019 | 106/4,993 | NR | 2.13 (1.73–2.55) |
| 2020 | 40/1,106 | NR | 3.63 (2.58–4.79) |
| 2021 | 55/1,040 | NR | 5.30 (3.98–6.70) |
| 2022 | 102/1,138 | NR | 8.99 (7.38–10.73) |
| 2023 | 89/1,056 | NR | 8.44 (6.71–10.22) |
| **Cardiorenal condition<sup>a</sup>** |  |  |  |
| ASCVD | 172/4,558 | 23 | 3.78 (3.22–4.34) |
| Heart failure | 142/2,676 | 22 | 5.32 (4.48–6.19) |
| CKD | 151/4,690 | 28 | 3.22 (2.73–3.74) |

Cumulative incidence was estimated with the Aalen–Johansen estimator, treating death before initiation as a competing event. 95% CIs are percentile intervals from 2,000 participant-level bootstrap samples. Index year is the calendar year in which follow-up began.
<sup>a</sup> Groups overlap; a participant may contribute to more than one group.
NR, not reported under the All of Us Data and Statistics Dissemination Policy; a protected death count does not by itself suppress the initiation estimate.
ASCVD, atherosclerotic cardiovascular disease; CI, confidence interval; CKD, chronic kidney disease; SGLT2, sodium-glucose cotransporter 2.

The 365-day cumulative incidence increased from 3.87% (95% CI 3.34–4.38) among participants entering the cohort in 2019 to 13.59% (95% CI 11.59–15.61) in 2022, and 12.25% (95% CI 10.29– 14.23) in 2023 (Table 2, Fig 2). Among the overlapping condition groups, the 365-day incidence was 8.40% (95% CI 7.41–9.53) for heart failure, 6.65% (95% CI 5.90–7.38) for ASCVD, and 5.28% (95% CI 4.62–5.92) for CKD.

**Fig 2.**
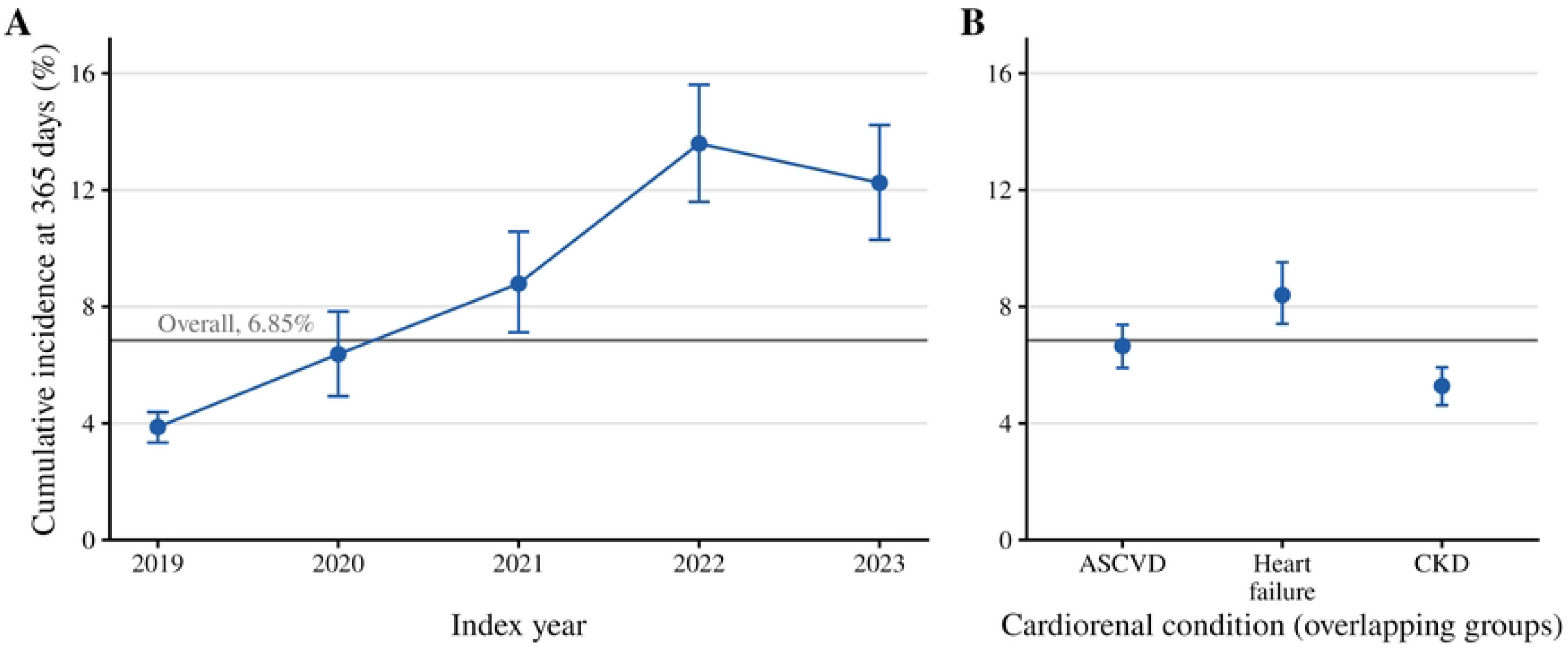
Cumulative incidence of recorded SGLT2 inhibitor initiation at 365 days by index year (A) and cardiorenal condition (B). Points are Aalen–Johansen estimates treating death before initiation as a competing event; error bars are 95% percentile bootstrap CIs. The horizontal line marks the overall estimate (6.85%). Condition groups overlap. ASCVD, atherosclerotic cardiovascular disease; CKD, chronic kidney disease; SGLT2, sodium-glucose cotransporter 2.

### Factors associated with initiation

Compared with participants entering the cohort in 2019, initiation was more likely among those entering in 2022 (adjusted risk ratio [aRR] 3.56, 95% CI 2.90–4.36) and 2023 (aRR 3.25, 95% CI

2.62–4.03). Heart failure was associated with higher initiation (aRR 1.51, 95% CI 1.29–1.76), whereas CKD was associated with lower initiation (aRR 0.63, 95% CI 0.54–0.74 No significant association was observed for ASCVD (aRR 1.04, 95% CI 0.89–1.21) (Table 3, Fig 3). Initiation did not differ significantly between participants with an eGFR of 45–59 (aRR 1.05, 95% CI 0.86– 1.28) or 30–44 mL/min/1.73 m² (aRR 1.11, 95% CI 0.87–1.41) and those with an eGFR ≥60 mL/min/1.73 m², but it was lower among individuals with an eGFR of 20–29 mL/min/1.73 m² (aRR 0.39, 95% CI 0.22–0.68). Relative to participants aged 45–64 years, initiation was less likely at ages 65–74 years (aRR 0.76, 95% CI 0.64–0.90) and ≥75 years (aRR 0.51, 95% CI 0.40–0.65).

**Fig 3.**
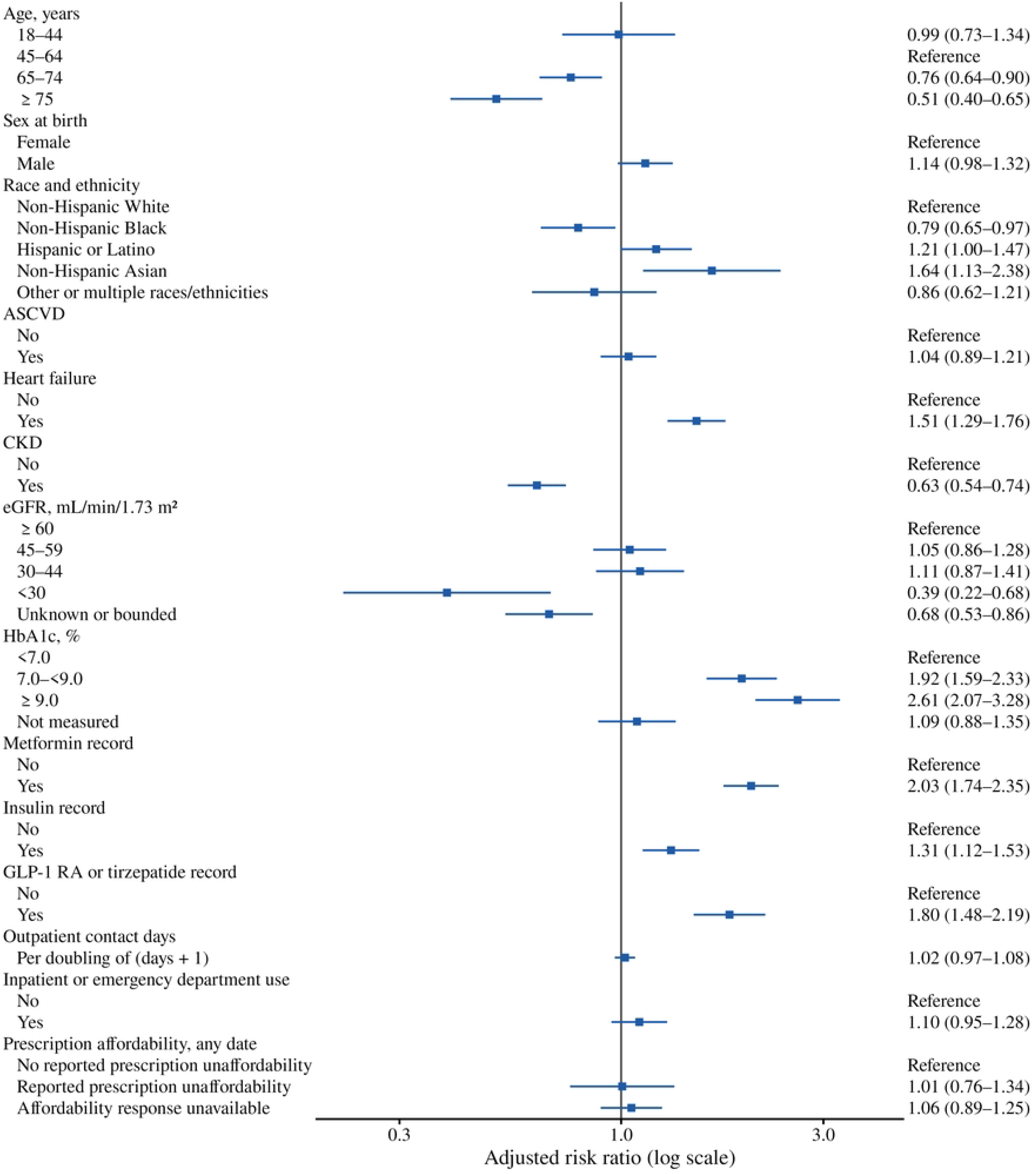
Adjusted associations of selected participant characteristics with recorded SGLT2 inhibitor initiation within 365 days. Each characteristic was modeled separately with modified Poisson regression adjusted for index year, age (natural cubic spline, df = 3), sex at birth, and race and ethnicity; and age was modeled in categories when it was the characteristic of interest. Risk ratios are not mutually adjusted. Estimates for all characteristics are shown in Table 3. ASCVD, atherosclerotic cardiovascular disease; CI, confidence interval; CKD, chronic kidney disease; eGFR, estimated glomerular filtration rate; GLP-1 RA, glucagon-like peptide-1 receptor agonist; HbA1c, glycated hemoglobin; RR, risk ratio.

**Table 3.** Associations of participant characteristics with recorded SGLT2 inhibitor initiation within 365 days.

| Characteristic | Unadjusted RR (95% CI) | Adjusted RR (95% CI) |
|---|---|---|
| **Index year** |  |  |
| 2019 | Reference | Reference |
| 2020 | 1.62 (1.24–2.12) | 1.56 (1.19–2.04) |
| 2021 | 2.26 (1.78–2.89) | 2.27 (1.78–2.90) |
| 2022 | 3.55 (2.90–4.35) | 3.56 (2.90–4.36) |
| 2023 | 3.13 (2.53–3.89) | 3.25 (2.62–4.03) |
| **Age, years** |  |  |
| 18–44 | 1.05 (0.77–1.43) | 0.99 (0.73–1.34) |
| 45–64 | Reference | Reference |
| 65–74 | 0.80 (0.67–0.95) | 0.76 (0.64–0.90) |
| ≥75 | 0.56 (0.43–0.72) | 0.51 (0.40–0.65) |
| **Sex at birth** |  |  |
| Female | Reference | Reference |
| Male | 1.11 (0.95–1.29) | 1.14 (0.98–1.32) |
| **Race and ethnicity** |  |  |
| Non-Hispanic White | Reference | Reference |
| Non-Hispanic Black | 0.79 (0.65–0.97) | 0.79 (0.65–0.97) |
| Hispanic or Latino | 1.19 (0.98–1.44) | 1.21 (1.00–1.47) |
| Non-Hispanic Asian | 1.55 (1.06–2.28) | 1.64 (1.13–2.38) |
| Other or multiple races/ethnicities | 0.88 (0.63–1.23) | 0.86 (0.62–1.21) |
| **ASCVD** |  |  |
| No | Reference | Reference |
| Yes | 0.97 (0.83–1.12) | 1.04 (0.89–1.21) |
| **Heart failure** |  |  |
| No | Reference | Reference |
| Yes | 1.37 (1.17–1.60) | 1.51 (1.29–1.76) |
| **CKD** |  |  |
| No | Reference | Reference |
| Yes | 0.62 (0.53–0.72) | 0.63 (0.54–0.74) |
| **eGFR, mL/min/1.73 m<sup>2</sup>** |  |  |
| ≥60 | Reference | Reference |
| 45–59 | 0.93 (0.76–1.13) | 1.05 (0.86–1.28) |
| 30–44 | 0.95 (0.75–1.21) | 1.11 (0.87–1.41) |
| 20–29 | NR | 0.39 (0.22–0.68) |
| Not measured or bounded | 0.67 (0.53–0.85) | 0.68 (0.53–0.86) |
| **HbA1c, %** |  |  |
| <7.0 | Reference | Reference |
| 7.0–<9.0 | 1.73 (1.43–2.10) | 1.92 (1.59–2.33) |
| ≥9.0 | 2.30 (1.83–2.88) | 2.61 (2.07–3.28) |
| Not measured | 1.05 (0.85–1.30) | 1.09 (0.88–1.35) |
| **Metformin record** |  |  |
| No | Reference | Reference |
| Yes | 2.08 (1.79–2.42) | 2.03 (1.74–2.35) |
| **Insulin record** |  |  |
| No | Reference | Reference |
| Yes | 1.26 (1.08–1.46) | 1.31 (1.12–1.53) |
| **GLP-1 RA or tirzepatide record** |  |  |
| No | Reference | Reference |
| Yes | 2.35 (1.97–2.81) | 1.80 (1.48–2.19) |
| **Outpatient contact days, per doubling of (days + 1)<sup>a</sup>** | 1.03 (0.97–1.09) | 1.02 (0.97–1.08) |
| **Inpatient or emergency department use** |  |  |
| No | Reference | Reference |
| Yes | 1.06 (0.91–1.23) | 1.10 (0.95–1.28) |
| **Difficulty affording prescriptions (any date)<sup>b</sup>** |  |  |
| No | Reference | Reference |
| Yes | 1.06 (0.80–1.40) | 1.01 (0.76–1.34) |
| No response | 0.92 (0.79–1.08) | 1.06 (0.89–1.25) |

Each characteristic was examined in separate modified Poisson models. Adjusted models included index year, age (natural cubic spline, df =3), sex at birth, and race and ethnicity; a characteristic was not adjusted for itself, and age was analyzed in categories when it was the characteristic of interest. Death before initiation was classified as no initiation; participants censored without an event before day 365 and those with unknown sex at birth or race and ethnicity were excluded.
<sup>a</sup> Outpatient contact days were modeled as log2(days + 1). Outpatient encounter days were defined as distinct calendar days with at least one recorded outpatient visit during the 365 days before the index date
<sup>b</sup> Survey responses from any date, which may precede or follow the index date.
NR, not reported under the All of Us Data and Statistics Dissemination Policy. Unadjusted estimates that could reveal a protected count are withheld; adjusted estimates are reported for modeled groups with more than 20 participants.
ASCVD, atherosclerotic cardiovascular disease; CI, confidence interval; CKD, chronic kidney disease; ED, emergency department; eGFR, estimated glomerular filtration rate; GLP-1 RA, glucagon-like peptide-1 receptor agonist; HbA1c, glycated hemoglobin; HC3, heteroskedasticity-consistent type 3; RR, risk ratio; SGLT2, sodium-glucose cotransporter 2.

Furthermore, non-Hispanic Black participants were less likely (aRR 0.79, 95% CI 0.65–0.97), and Asian participants were more likely (aRR 1.64, 95% CI 1.13–2.38), to initiate treatment compared to non-Hispanic White participants. Initiation was more likely among patients with an HbA1c of 7.0% to <9.0% (aRR 1.92, 95% CI 1.59–2.33) and ≥9.0% (aRR 2.61, 95% CI 2.07–3.28) compared with an HbA1c <7.0%. Initiation was also higher among those with baseline records of metformin (aRR 2.03, 95% CI 1.74–2.35), a glucagon-like peptide-1 receptor agonist (GLP-1 RA) or tirzepatide (aRR 1.80, 95% CI 1.48–2.19), or insulin (aRR 1.31, 95% CI 1.12–1.53). Finally, sex at birth, outpatient contact frequency, inpatient or emergency department use, and reported difficulty affording prescriptions were not significantly associated with initiation.

### Sensitivity analyses

Using calendar-specific eGFR thresholds, the resulting cohort included 9,257 participants, with a 365-day cumulative incidence of 6.97% (95% CI 6.48–7.47; Table D in S1 Appendix). In the mutually adjusted sensitivity models, associations of heart failure, CKD, index year, HbA1c, metformin, and GLP-1 RA or tirzepatide with initiation remained generally consistent across analyses (S1 Appendix). When applying inverse probability of censoring weights, the aRRs were 1.41 (95% CI 1.17–1.70) for heart failure and 0.62 (95% CI 0.50–0.78) for CKD; after excluding index-day medication records, the corresponding aRRs were 1.44 (95% CI 1.20–1.74) and 0.63 (95% CI 0.50–0.78).

## Discussion

Among 9,333 adults in the All of Us Research Program with T2DM and ASCVD, heart failure, or CKD who had no prior recorded SGLT2 inhibitor use, the estimated 365-day cumulative incidence of recorded initiation was 6.85%. Incidence was higher in 2022 (13.59%) and 2023 (12.25%) than in 2019 (3.87%). Initiation was more likely among patients with heart failure or higher HbA1c, and less likely among those with CKD, older adults, and non-Hispanic Black participants.

Our findings reflect a different measure of SGLT2 inhibitor utilization than those reported in previous studies. Previous studies, including earlier All of Us analyses [20,21], reported prevalent or ever-recorded use rather than first initiation during a defined period; therefore, contemporary estimates of 12%–20% [15–17] are not directly comparable with the 1-year cumulative incidence of initiation among participants without prior recorded use. One U.S. EHR study reported that 7.0% of patients with a new ASCVD diagnosis in 2022 initiated an SGLT2 inhibitor alone within 1 year [22], although differences in study populations, eligibility criteria, and medication ascertainment limit direct comparison.

The increase across entry years coincided with growing evidence of SGLT2 inhibitor benefits in heart failure [37–40] and CKD [41,42], along with broader guideline recommendations [4–8]. Calendar year also reflects changes in labeling, health care coverage, cost, and clinician familiarity, which this design cannot investigate separately. Because each participant entered on their first eligible date, entry-year groups may differ in composition; differences between years reflect both calendar time and who became eligible in each year. Higher initiation among participants with heart failure is consistent with the rapid incorporation of SGLT2 inhibitors into guideline-directed heart failure therapy [6]. Even so, the 365-day cumulative incidence was 8.40% among participants with heart failure, in line with registry evidence of persistent underuse [17]. CKD was associated with lower initiation despite trial evidence of kidney and cardiovascular benefit [3,41,42] and expanding guideline recommendations [5,8], consistent with findings in Veterans Affairs and health-system cohorts [18,19]. SGLT2 inhibitor initiation declined with age, consistent with previous nationwide cohort data on early adoption [13]. The relative benefit of dapagliflozin in the DAPA-HF trial was consistent in patients aged ≥75 years [40]. However, clinicians may reasonably consider volume depletion, genital infections, polypharmacy, frailty, and life expectancy when prescribing these medications; therefore, this decrease in initiation among older adults may be partially explained by individualized patient care. Non-Hispanic Black participants were less likely than non-Hispanic White participants to initiate treatment. Similar differences have been reported among commercially insured patients, veterans, and patients with CKD, persisting after extensive clinical and socioeconomic adjustment [14,19,44]. This association may reflect a combination of clinical characteristics, care settings, and access barriers that these data cannot distinguish.

Initiation was more likely among participants with higher HbA1c, although guidelines in effect during the study recommended SGLT2 inhibitors for cardiorenal risk reduction regardless of HbA1c [4,7]. This pattern suggests that initiation may have remained influenced by the need for glycemic intensification. Larger treatment gaps have been reported among patients with well-controlled diabetes and kidney disease [45], and Veterans Affairs patients receiving SGLT2 inhibitors had higher HbA1c levels than non-recipients [16]. Whether lower recorded initiation in these groups reflects appropriate clinical decisions, access barriers, or incomplete capture of prescriptions requires further study. Targeted EHR alerts modestly increased guideline-directed prescribing in heart failure in a cluster-randomized trial [46], and similar approaches could be evaluated for cardiorenal indications in diabetes.

### Strengths and limitations

Strengths of this study include its large, diverse cohort and the estimation of first recorded SGLT2 inhibitor initiation among adults with T2DM and established cardiorenal disease. We identified a single index date for each participant and estimated the cumulative incidence of initiation within 365 days. The 2019–2023 study period captured changes in treatment uptake during a period of expanding clinical evidence and guideline recommendations.

Our findings should be interpreted in the context of several limitations. First, All of Us participants are volunteers, and EHR completeness varies across contributing organizations [26]. Furthermore, the requirement for one year of prior EHR activity may preferentially select participants engaged in regular care; therefore, our estimates are not nationally representative. Second, the outcome was based on a medication record rather than a dispensing record. Prescriptions written outside contributing health systems may have been missed, which could underestimate true initiation rates and misclassify prevalent users as new users. Third, the phenotype algorithms were adapted from claims-based definitions and have not been validated against chart review within the All of Us cohort. The requirement for repeated outpatient diagnoses may have excluded patients with genuine T2DM, heart failure, CKD, or ASCVD documented only once, particularly when care occurred outside contributing health systems. In addition, diagnosis-based CKD ascertainment may have missed cases identifiable through laboratory findings alone. Fourth, the eGFR threshold of 20 mL/min/1.73 m² was applied uniformly across study years despite evolving recommendations. Because participants without a recent eGFR were retained, baseline kidney-function eligibility was unknown for some individuals. Fifth, the index date marks when study criteria were met, not necessarily when treatment first became clinically indicated. Some participants may have had unrecorded contraindications or preferences that made non-initiation appropriate; thus, non-initiation should not strictly be equated with guideline-discordant care.

### Conclusions

Among adults with T2DM and established cardiorenal disease in the All of Us Research Program who had no prior recorded SGLT2 inhibitor exposure, fewer than 1 in 10 had a recorded initiation within 1 year. Although initiation increased from 2019 to 2023, overall rates remained suboptimal, particularly among participants with CKD, older adults, and non-Hispanic Black adults. These findings highlight a persistent gap between clinical evidence and real-world practice, indicating the need for targeted interventions to ensure equitable access and to promote SGLT2 inhibitor use for cardiorenal protection, independent of glycemic control.

## Data Availability

The data underlying this study are from the All of Us Research Program and are subject to third-party restrictions. Under the All of Us Data User Code of Conduct and data use agreement, the authors are not permitted to publicly share or redistribute participant-level data. The data are available to qualified researchers who complete All of Us registration, training, and an institutional data use agreement, via the All of Us Researcher Workbench (https://www.researchallofus.org). This study used the Controlled Tier Curated Data Repository version 9 (CDR v9, release C2025Q4R6). The authors had no special access privileges that other qualified researchers would not have. Concept-set definitions required to reproduce the study cohort are provided in the S1 Appendix.

## Acknowledgments

We gratefully acknowledge the All of Us participants, without whom this research would not have been possible. We also thank the National Institutes of Health’s All of Us Research Program for making the participant data examined in this study available.

## Supporting information

S1 Appendix. Supplementary methods and results. Phenotype and laboratory definitions (Table A), Participant counts by confirmation route (Table B), characteristics by cardiorenal condition (Table C), sensitivity analyses (Tables D and E), and timing of survey responses (Table F).

**S1 Checklist. STROBE and RECORD checklists.** Completed STROBE checklist for cohort studies and RECORD extension checklist with manuscript locations.

